# Critical Gaps in Nutritional Care for Adolescents Living with HIV: A Multi-Facility Study from Ethiopia

**DOI:** 10.1101/2025.08.01.25332677

**Authors:** Meless Gebrie Bore, Lin Perry, Xiaoyue Xu, Andargachew Kassa Biratu, Marilyn Cruickshank

## Abstract

**Background:** Adolescents living with HIV (ALHIV) in Ethiopia face significant nutritional challenges affecting their health and ART outcomes. Increased energy needs, HIV complications, and socioeconomic barriers contribute to their vulnerability. Despite ART advancements, research on their nutritional care remains limited, and comprehensive guidance is lacking. This study assessed ALHIV’s nutritional status, nutrition support in ART clinics, and strategies for improvement.

**Method:** A two cross-sectional survey were conducted in ten public hospitals across Addis Ababa and Oromia regions, involving 384 ALHIV and 44 healthcare professionals. Healthcare workers were selected through purposive sampling, while ALHIV were recruited using proportionate random sampling. Data was collected using a pre-tested structured questionnaire with quantitative and qualitative components, administered by trained healthcare workers via the Kobo Toolbox program.

**Results:** Nutritional assessments of ALHIV revealed that 24.2% were thin, 21.7% were stunted, and 34.9% were malnourished based on mid-upper arm circumference, with 19.4% experiencing severe acute malnutrition. Many adolescents faced food insecurity, which negatively affected their nutritional status and ART adherence.

While most healthcare workers conducted basic nutritional assessments, the use of sensitive tools was limited. Only 36.4% assessed dietary intake, 27.3% evaluated food security, and 38.6% provided regular nutrition counseling. Healthcare professionals expressed dissatisfaction with the integration of nutrition services due to inadequate training and resource constraints.

**Conclusion:** The study highlights significant gaps in nutrition support and malnutrition among ALHIV on ART in Ethiopia. Integrating nutritional assessments and counseling into routine ART care, enhancing healthcare worker training, and developing standardized nutritional guidelines are essential for improving outcomes. Addressing food insecurity and socioeconomic barriers through adequate resources and policies is also critical for better health outcomes. Further research is needed to understand the long-term nutritional needs of ALHIV.

## 1. Introduction

Adolescents living with HIV (ALHIV) face a dual burden of managing a chronic infection during a life stage characterized by rapid physical, emotional, and cognitive development^[1]^. In Ethiopia, where adolescents represent approximately 33% of the population ^[2]^, this group is particularly vulnerable to malnutrition due to increased nutritional needs, the physiological impacts of HIV, and widespread food insecurity. Malnutrition among ALHIV undermines immune function, increases susceptibility to infections, and impairs treatment outcomes, presenting a critical challenge to adolescent health and HIV care systems^[3]^.

Adequate nutrition is essential for optimal antiretroviral therapy (ART) effectiveness and long-term health outcomes among adolescents ^[4, 5]^. HIV further elevates energy and nutrient requirements, and poor nutritional status in adolescence can impair growth, delay puberty, and increase the risk of non-communicable diseases in adulthood ^[5–7]^. Studies have documented that ALHIV commonly experience undernutrition, micronutrient deficiencies, and, in some contexts, overweight or obesity^[3]^. In Ethiopia, these challenges are intensified by food insecurity, limited healthcare infrastructure, and inadequate integration of nutritional services into ART care ^[8, 9]^. Factors such as peer pressure, household income, parental influence, and cultural beliefs also complicate adolescent dietary behaviors and adherence to nutritional guidance ^[10]^.

While the role of nutrition in the management of HIV is well-established, there is limited evidence from Ethiopia on the actual nutritional status of ALHIV and the practices of healthcare providers in assessing and addressing their nutritional needs. Specifically, few studies have examined how healthcare workers assess nutritional status, provide counselling, or implement support services in ART settings. Moreover, there is a lack of data on the tools, training, and resources available to healthcare workers to deliver comprehensive nutritional care for ALHIV.

This study aimed to assess the anthropometric and nutritional status of adolescents receiving ART in Ethiopia and to explore the nutritional assessment, counselling, and support practices of healthcare professionals working in ART clinics. By identifying gaps in service delivery and highlighting areas for improvement, the study seeks to inform evidence-based strategies and policies to strengthen nutritional care and improve health outcomes for adolescents living with HIV.

## 2. Methods

### 2.1. Study Setting and period

This study was conducted between August and December 2023 across two regions in Ethiopia: Addis Ababa and Oromia. Ten hospitals were purposively selected based on their high HIV burden and large number of ART recipients: Adama Referral Hospital, ALERT General Hospital, Asella Referral and Teaching Hospital, Batu Hospital, Bishoftu Hospital, Ras Desta Damtew Hospital, Shashamene Comprehensive Specialized Hospital, St Paul’s Comprehensive Specialized Hospital, Yekatit 12 Hospital, and Zewditu Hospital.

### 2.2. Study Design and population

An institution-based, cross-sectional survey design was employed, using both quantitative and qualitative components. The quantitative survey assessed the nutritional status of ALHIV and evaluated healthcare workers’ practices related to nutritional assessment, counselling, and management. The study adhered to the Strengthening the Reporting of Observational Studies in Epidemiology – Nutritional Epidemiology (STROBE-Nut) guidelines.^[11–13]^.

### 2.3. Sampling and recruitment

#### 2.3.1. Sampling of ALHIV

A proportionate random sampling strategy was used to recruit ALHIV aged 10–19 years based on data provided by ART data clerks at each facility. A sampling frame was developed based on ART registration codes. Participants were assigned unique research codes to ensure confidentiality.

Sample sizes were allocated proportionally according to the number of ALHIV in each hospital. Within each hospital, eligible participants were randomly selected using SPSS version 26 based on their registration numbers.

### ALHIV Inclusion Criteria

- Aged 10–19 years
- Currently on ART
- Receiving care at one of the selected hospitals

### ALHIV Exclusion Criteria

- On ART for less than three months
- Cognitive or communication impairments preventing reliable participation
- Under 18 years of age without guardian/parental consent
- Missing ART registration number or unable to verify identity

#### 2.3.2. Sampling of Healthcare Professionals

- A purposive sampling method was used to select healthcare professionals at each study site. These participants contributed qualitative data regarding their practices in nutritional assessment and support.

### Inclusion Criteria

- Actively involved in delivering ART to ALHIV at the selected facilities

### Exclusion Criteria

- Employed in the ART unit for less than three months

### 2.4. Sample Size Determination

#### 2.4.1. ALHIV Sample Size

The sample size for ALHIV was determined based on three primary objectives: assessing nutritional status, identifying influencing factors, and evaluating food consumption patterns. The largest required sample size across these objectives was selected to ensure adequate power.

- **Nutritional status:** Using a single population proportion formula (5% margin of error, 95% CI, and a 33.1% undernutrition prevalence from a 2020 study in southern Ethiopia ^[14]^), the required sample size was 340 participants.
- **Influencing factors:** A two-population proportion formula (5% type I error, 80% power, 1:1 ratio) was used to calculate a sample size of 352 participants for the variable “meal skipping” ^[14–16]^.
- **Food consumption patterns:** Assuming a 50% prevalence (due to lack of prior data), the estimated sample size was 384 participants.

The final sample size of 384 ALHIV was selected to satisfy all study objectives.

#### 2.4.2 Healthcare Professional Sample Size

A total of 50 healthcare professionals were identified across the ten hospitals (approximately five per site). Of these, four were unavailable due to annual leave (n=1), offsite training (n=2), or maternity leave (n=1). Among the 46 invited participants, 44 consented to participate.

### 2.5. Data Collection

Data were collected using a pre-tested, interviewer-administered structured questionnaire, which included both closed-and open-ended questions tailored for ALHIV and healthcare professionals. The Kobo Toolbox platform ^[17]^ was used to facilitate electronic data collection via smartphones and computers. Interviews were conducted by trained healthcare professionals.

A three-day training workshop was held prior to data collection to prepare the data collectors. The training covered the study objectives, data collection tools, interview techniques, and procedures for nutritional assessments, including anthropometric measurements, clinical examinations, and dietary intake assessments.

Clinical assessments followed standardized protocols (see Supplementary File Box 1) to ensure consistency and accuracy. Measurements were performed using calibrated instruments, including digital scales and portable stadiometers ^[18]^. Physical examinations included the assessment of signs of malnutrition, such as wasting and pallor. To verify the reliability of anthropometric measurements, the Intraclass Correlation Coefficient (ICC) was calculated, yielding a value of 0.856, indicating good measurement agreement ^[19, 20]^.

### 2.6. Survey Questionnaires

The questionnaires were initially developed in English and translated into Amharic. To ensure linguistic accuracy, the Amharic version was back-translated into English by a language expert, and both versions were compared for consistency ^[21]^.

Content validity was assessed using the Item-Level Content Validity Index (I-CVI) and the Scale-Level Content Validity Index (S-CVI) ^[22, 23]^, with a threshold of 0.78 or higher indicating acceptable relevance ^[24]^.

Inter-rater consistency was evaluated using Cohen’s Kappa coefficient, which achieved a value of 0.71, indicating substantial agreement ^[19]^. Qualitative feedback from the expert panel was incorporated to improve clarity and refine the questionnaire.

### 2.7. Data Processing and Analysis

All quantitative data were analysed using SPSS version 21. Descriptive statistics were generated to examine nutrition-related assessment, counselling, and management practices. Frequencies, percentages, and measures of central tendency were calculated, with results presented in tables and figures. Data were screened for outliers and inconsistencies.

Normality of continuous variables was assessed using both the Kolmogorov-Smirnov and Shapiro-Wilk tests (significance threshold p < 0.05), and visually validated through Q-Q plots. To identify associations among anthropometric indices, correlational and linear regression analyses were conducted.

Open-ended qualitative responses were coded and categorized into thematic variables for integration with the quantitative dataset. These coded data were analysed alongside quantitative findings to offer deeper insights into healthcare professionals’ practices. Participants were identified by study codes to maintain confidentiality.

Operational definitions used in the study are provided in Supplementary File Box 2.

### 2.8. Ethics Approval

This study was conducted in accordance with the Declaration of Helsinki. Ethical approval was obtained from the Human Research Ethics Committee at the University of Technology Sydney, Australia (Ref. Number: ETH23-7873); the Institutional Review Board of the College of Medicine and Health Sciences, Hawassa University, Ethiopia (Ref. Number: IRB/321/15) — which also covered the Oromia region study sites; the Addis Ababa City Administration Health Bureau Ethics Review Committee, Ethiopia (Ref No: A/A/3/54/227); and St. Paul’s Hospital Millennium Medical College Institutional Review Board (Ref No: PM23/181).

Permission was secured from all health institutions involved in the study. Written informed consent for participation was obtained from participants’ legal guardians or next of kin. Confidentiality was assured and maintained throughout the study

## 3. Results

### 3.1. Participants’ characteristics

#### Adolescents Living with HIV (ALHIV)

A total of 384 adolescents living with HIV (ALHIV) receiving ART follow-up services participated in the study, yielding a 100% response rate. The mean age of participants was 15.9 ± 2.19 years, with the majority (n = 227; 59.1%) aged between 14 and 17 years, representing the middle adolescent age group. More than half were female (n = 207; 54%). Nearly all participants were students (n = 379; 98.7%), with close to half (n = 179; 46.6%) enrolled in grades 1 to 8.

In terms of household characteristics, the largest proportion of participants came from households earning between 1,000 and 3,000 Ethiopian Birr (EBR) per month (n = 175; 45.6%). More than half (n = 198; 51.6%) lived in households with 4–5 members (Table 2).

**Table 1a.**
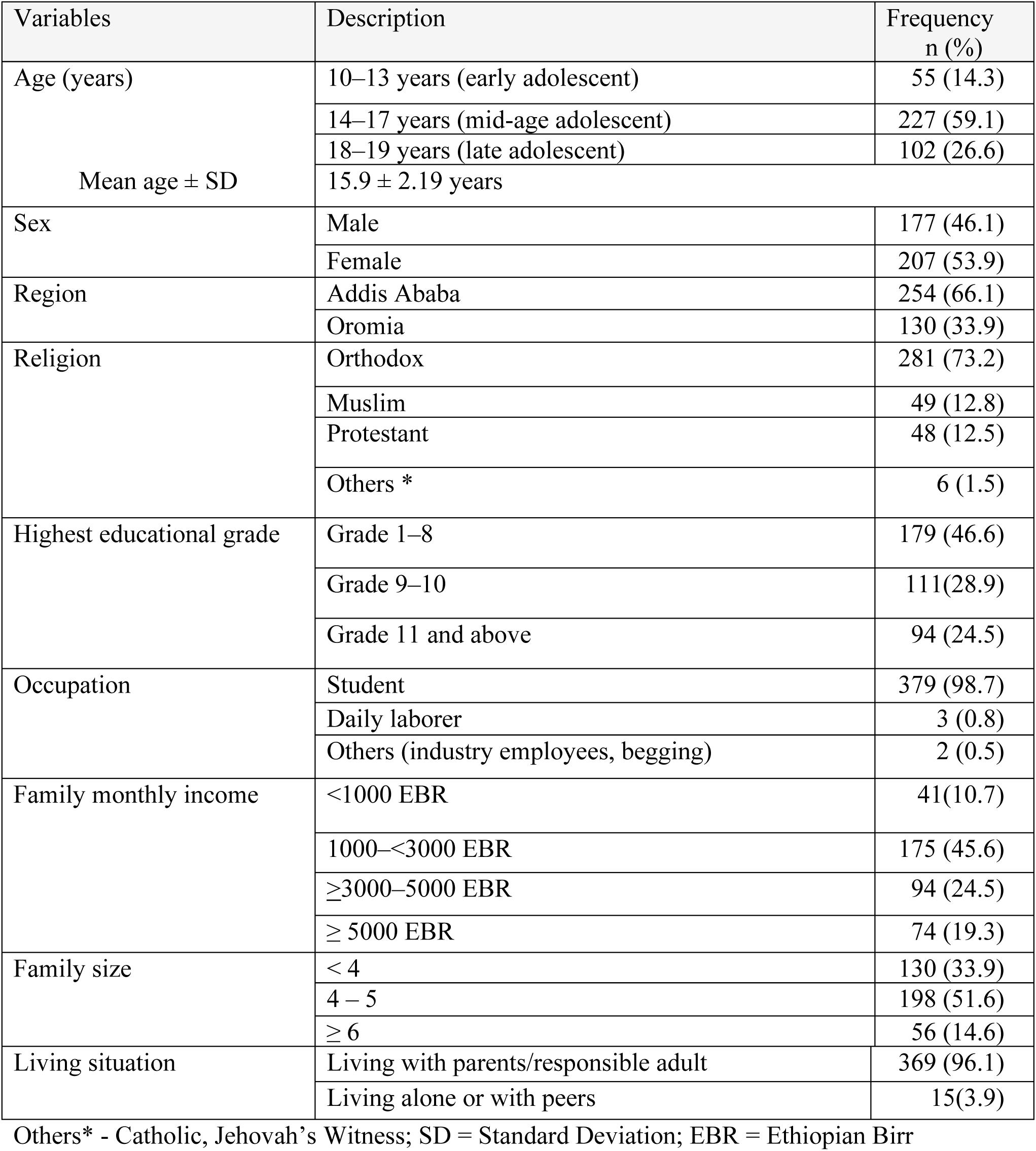
Socio-demographic characteristics of 384 ALHIV attending ART follow-up across 10 selected hospitals in Addis Ababa and Oromia regions, Ethiopia.

| Variables | Description | Frequency<br>n (%) |
| --- | --- | --- |
| Age (years) | 10–13 years (early adolescent) | 55 (14.3) |
|  | 14–17 years (mid-age adolescent) | 227 (59.1) |
|  | 18–19 years (late adolescent) | 102 (26.6) |
| | Mean age ± SD | 15.9 ± 2.19 years |
| Sex | Male | 177 (46.1) |
|  | Female | 207 (53.9) |
| Region | Addis Ababa | 254 (66.1) |
|  | Oromia | 130 (33.9) |
| Religion | Orthodox | 281 (73.2) |
|  | Muslim | 49 (12.8) |
|  | Protestant | 48 (12.5) |
|  | Others * | 6 (1.5) |
| Highest educational grade | Grade 1–8 | 179 (46.6) |
|  | Grade 9–10 | 111(28.9) |
|  | Grade 11 and above | 94 (24.5) |
| Occupation | Student | 379 (98.7) |
|  | Daily laborer | 3 (0.8) |
|  | Others (industry employees, begging) | 2 (0.5) |
| Family monthly income | <1000 EBR | 41(10.7) |
|  | 1000–<3000 EBR | 175 (45.6) |
| | ≥3000–5000 EBR | 94 (24.5) |
| | ≥ 5000 EBR | 74 (19.3) |
| Family size | < 4 | 130 (33.9) |
|  | 4 – 5 | 198 (51.6) |
| | ≥ 6 | 56 (14.6) |
| Living situation | Living with parents/responsible adult | 369 (96.1) |
|  | Living alone or with peers | 15(3.9) |

Others\* - Catholic, Jehovah's Witness; SD = Standard Deviation; EBR = Ethiopian Birr

**Table 1b.** Socio-Demographic Characteristics of the ART Clinic Healthcare Workers in the Selected Hospitals of two Regions of Ethiopia, 2024.

| Variables | Description | Frequency<br>N (%) |
| --- | --- | --- |
| Age (year) | 24 – 34 years | 21(47.7) |
|  | 35 – 44 years | 16 (36.4) |
|  | ≥ 45 years | 7 (15.9) |
|  | Mean ± SD =35.4 ± 6.99 year |  |
| Sex | Male | 17 (38.6) |
|  | Female | 27 (61.4) |
| Region | Addis Ababa | 25 (56.8) |
|  | Oromia | 19 (43.2) |
| Educational Status | Diploma Graduate | 4 (9.1) |
|  | Bachelor's Degree Graduate | 34 (77.3) |
|  | Master's Degree Graduate | 6 (13.6) |
| Profession | Registered Nurse | 28 (63.6) |
|  | Public Health Officer | 6 (13.6) |
|  | General Practitioner | 10 (22.7) |
| Length of Work Experience | < 10 years | 26 (59.1) |
|  | 10 – 20 years | 14 (31.8) |
|  | ≥ 21 years | 4 (9.1) |
|  | Mean ± SD = 10.7 ± 6.61 year |  |
| Length of Work Experience in<br>ART unit/clinic | < 10 years | 36 (81.9) |
|  | ≥ 10 years | 8 (18.2) |
|  | Mean ± SD = 5.45 ± 4.23 year |  |
| Monthly Income | 5000 – 7500 EBR | 9 (20.5) |
|  | 7501 – 10000 EBR | 19 (43.2) |
|  | > 10000 EBR | 16 (36.4) |

SD - Standard Deviation

**Table 2.**
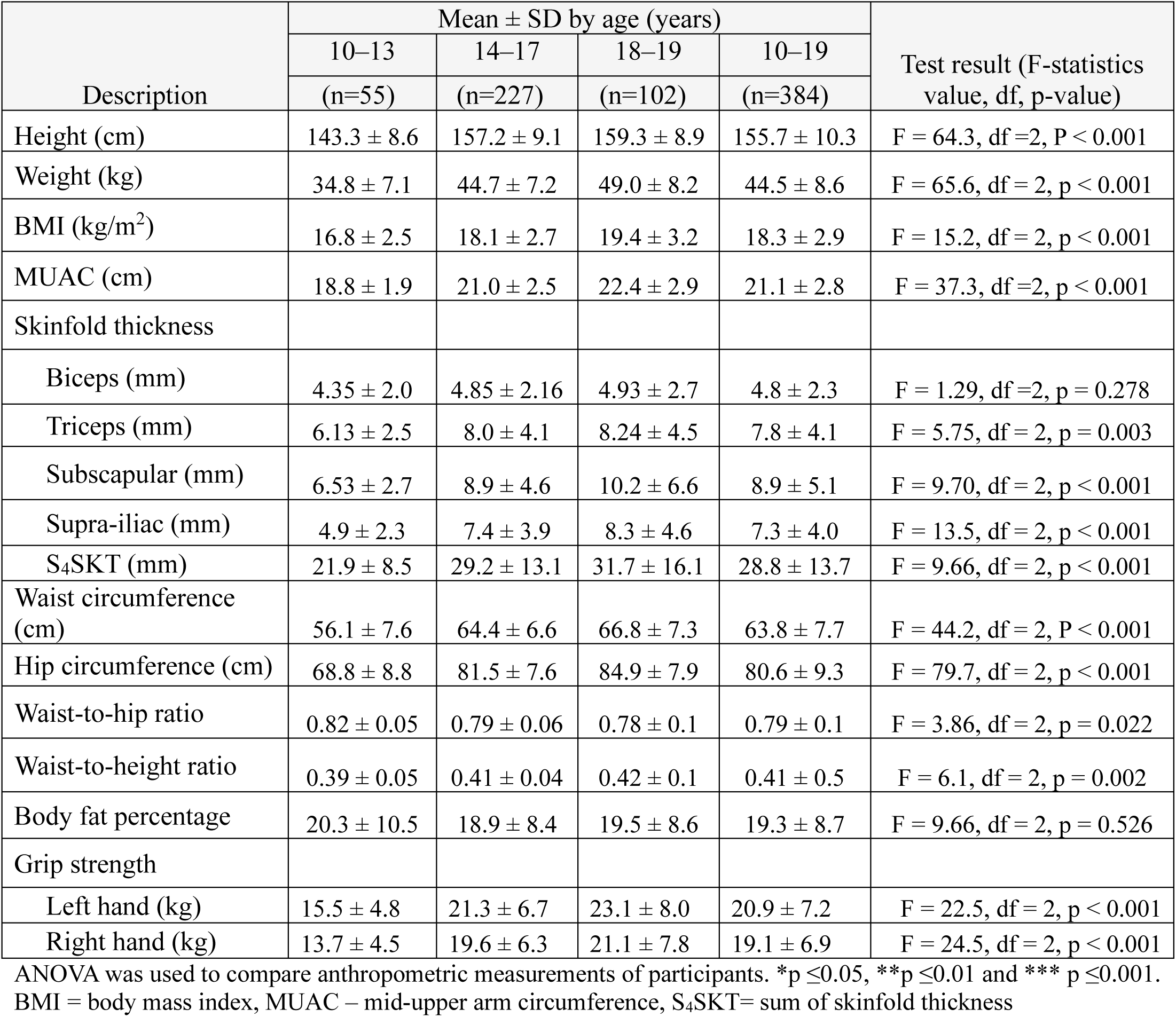
Descriptive analysis of ALHIVs’ anthropometric variables by age category.

#### Healthcare Professionals

A total of 44 healthcare workers participated, reflecting a 95.6% response rate. The mean age of respondents was 35.4 ± 6.99 years, with nearly half (n = 21; 47.7%) aged 24–34 years. More than half were recruited from Addis Ababa regional hospitals (n = 25; 56.8%).

In terms of professional background, most respondents held bachelor’s degrees (n = 34; 77.3%) and were Registered Nurses (n = 28; 63.6%). The majority (n = 26; 59.1%) had less than 10 years of professional experience, and most reported previous work in an ART clinic (n = 36; 81.9%) (Table 3).

**Table 3.** Pearson correlation analysis for anthropometric indices.

| Variables | BFP | BMI-for-Age (BAZ) | MUAC-for-age (cm) | S4SKT (mm) |
| --- | --- | --- | --- | --- |
| BFP | - | 0.264** | 0.148** | 0.436** |
| BMI-for-Age (BAZ) | 0.264** | - | 0.377** | 0.335** |
| MUAC-for-age | 0.148** | 0.377** | - | 0.298** |
| S4SKT | 0.436** | 0.335** | 0.298** | - |

Data presented are correlation coefficient (r) values.
Bold values indicate significant positive correlations.
\*\* p < 0.001 (2-tailed)
BFP = body fat percentage; BMI = body mass index; MUAC = mid-upper arm circumference;
S4SKT = the sum of four skinfold thicknesses (biceps, triceps, subscapular, and supra-iliac SKT)

### 3.2. Anthropometric and Nutritional Status of Adolescents Living with HIV

#### 3.2.1. Anthropometric Measurements

The mean height and weight of ALHIV participants were 155.7 ± 10.3 cm and 44.5 ± 8.6 kg, respectively. The mean body mass index (BMI) was 16.8 ± 2.5 kg/m², and the mean mid-upper arm circumference (MUAC) was 21.0 ± 2.8 cm. Statistically significant differences in anthropometric parameters were observed across different age groups (Table 4).

**Table 4.** Anthropometric measurements associated with thinness among ALHIV on ART follow-up.

| Variable | Thinness (undernutrition with BMI-for-Age Z-score indicators) |  |
| --- | --- | --- |
|  | Simple LR | Adjusted LR |
| | $\beta$ coefficient (95%CI) | $\beta$ coefficient (95% CI) |
| MUAC < 21 cm | -0.38 (-0.42, -0.26) *** | -0.37 (-0.42, -0.26) *** |
| (S4SKT < 25 cm | -0.34 (-0.37, -0.21) *** | -0.25 (-0.29, -0.13) *** |
| Hip circumference < 82 cm | -0.33 (-0.37, -0.20) *** | -0.16 (-0.23, -0.05) ** |
| Waist circumference < 64 cm | -0.21 (-0.26, -0.10) *** |  |

Significant at \*P value $\leq 0.05$ , \*\*P value $\leq 0.01$ and p\*\*\* value $\leq 0.001$
Model fitness –
F-statistics – 35.09, PV < 0.001 Adjusted R<sup>2</sup>=0.211; SE=0.381; Durbin Watson=1.935
Model 1 - Predictors MUAC
Model 2 -Predictors: MUAC, S4SKT
Model 3 -Predictors: MUAC, S4SKT, and hip circumference
Dependent Variable: Thinnes growth

#### 3.2.2. Nutritional Status

Nutritional status was assessed using three indicators: BMI-for-Age Z-score (BAZ) to determine thinness, Height-for-Age Z-score (HAZ) for stunting, and MUAC-for-age for acute malnutrition.

### **a)** Thinness Based on BMI-for-Age

Using the 2007 WHO Growth Reference ^[25]^, 24.2% (n = 93) of ALHIV were classified as thin (BAZ <-2 SD), of whom 26.9% (n = 25) were severely thin (BAZ <-3 SD) and 73.1% (n = 68) moderately thin (-3 SD ≤ BAZ <-2 SD) ^[25–27]^ (Figure 1a). Thinness was more prevalent among males than females. The highest prevalence of thinness was observed in late adolescence (Figure 1b).

**Figure 1a.**
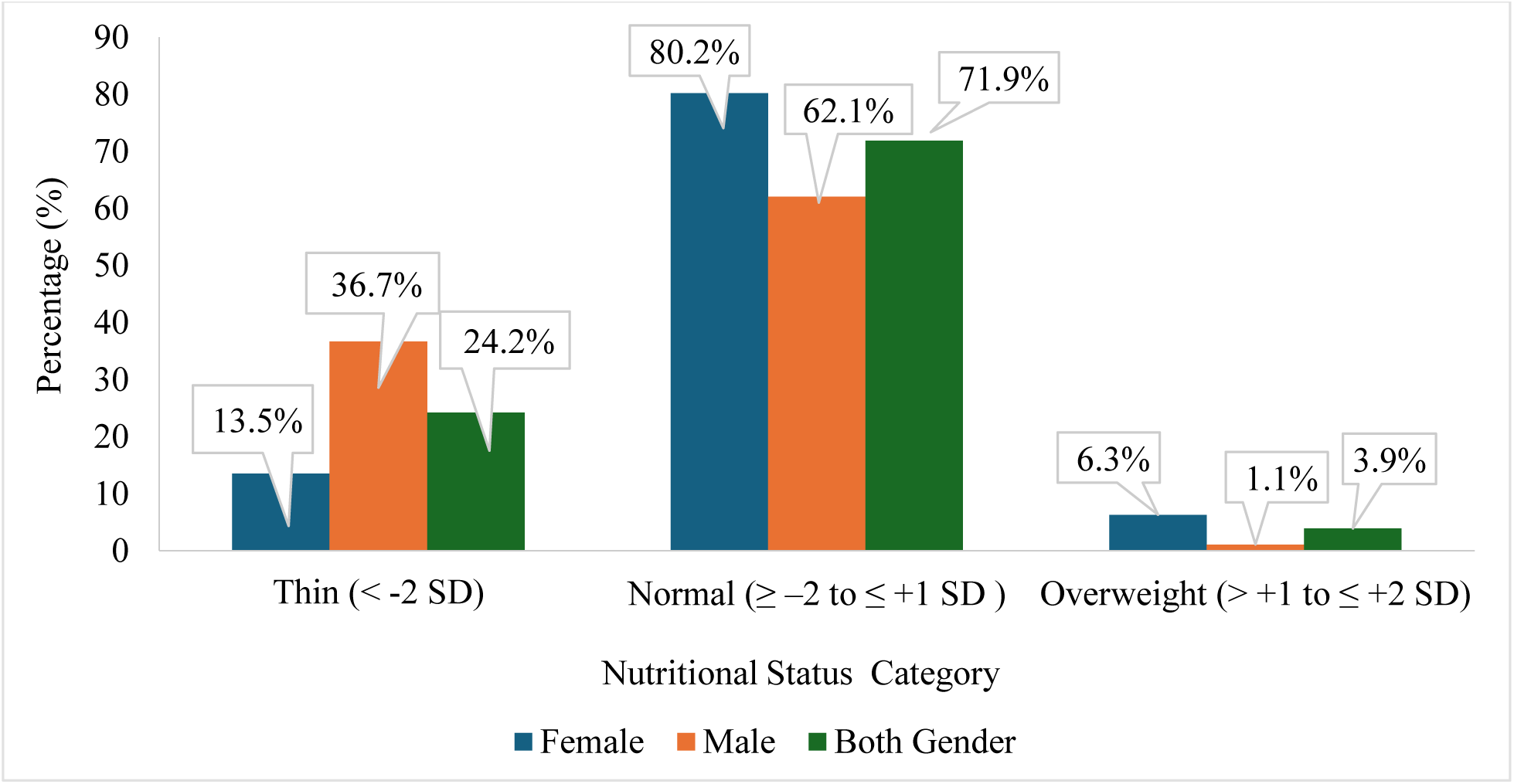
Level of undernutrition assessed as BMI-for-Age by gender of ALHIV on ART follow-up.

**Figure 1b.**
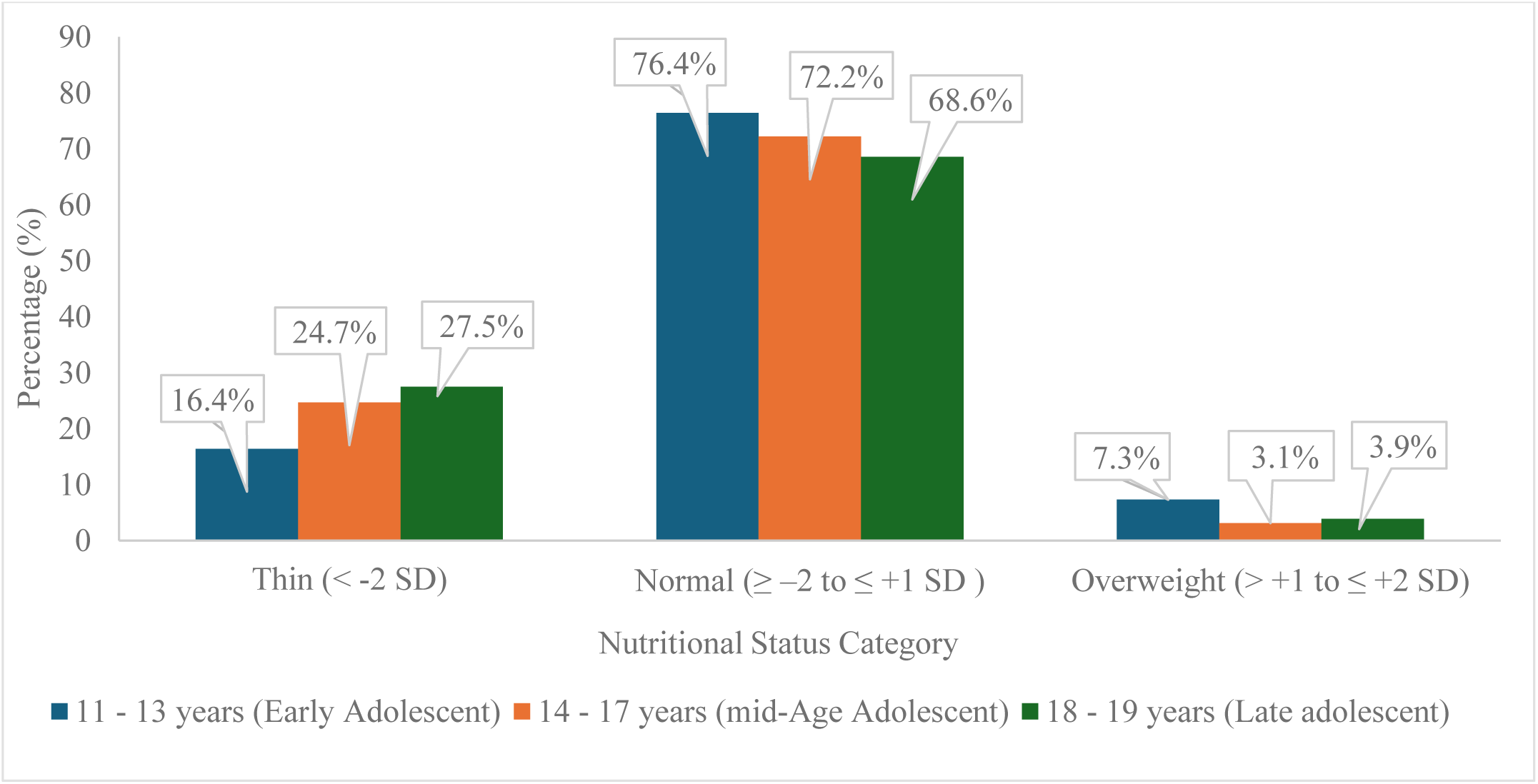
Distribution of nutritional status according to BMI-for-age by age of ALHIV on ART follow-up.

### **b)** Stunting Based on Height-for-Age

A total of 21.7% (n = 83) of participants were stunted (HAZ <-2 SD), with 28.9% (n = 24) of these classified as severely stunted (HAZ <-3 SD) ^[25]^. Severe stunting was most common among late adolescents (8.8%). Stunting prevalence was slightly higher in males (22.6%) compared to females (20.8%), with severe stunting in 7.3% of males and 5.3% of females (Figure 2).

**Figure 2.**
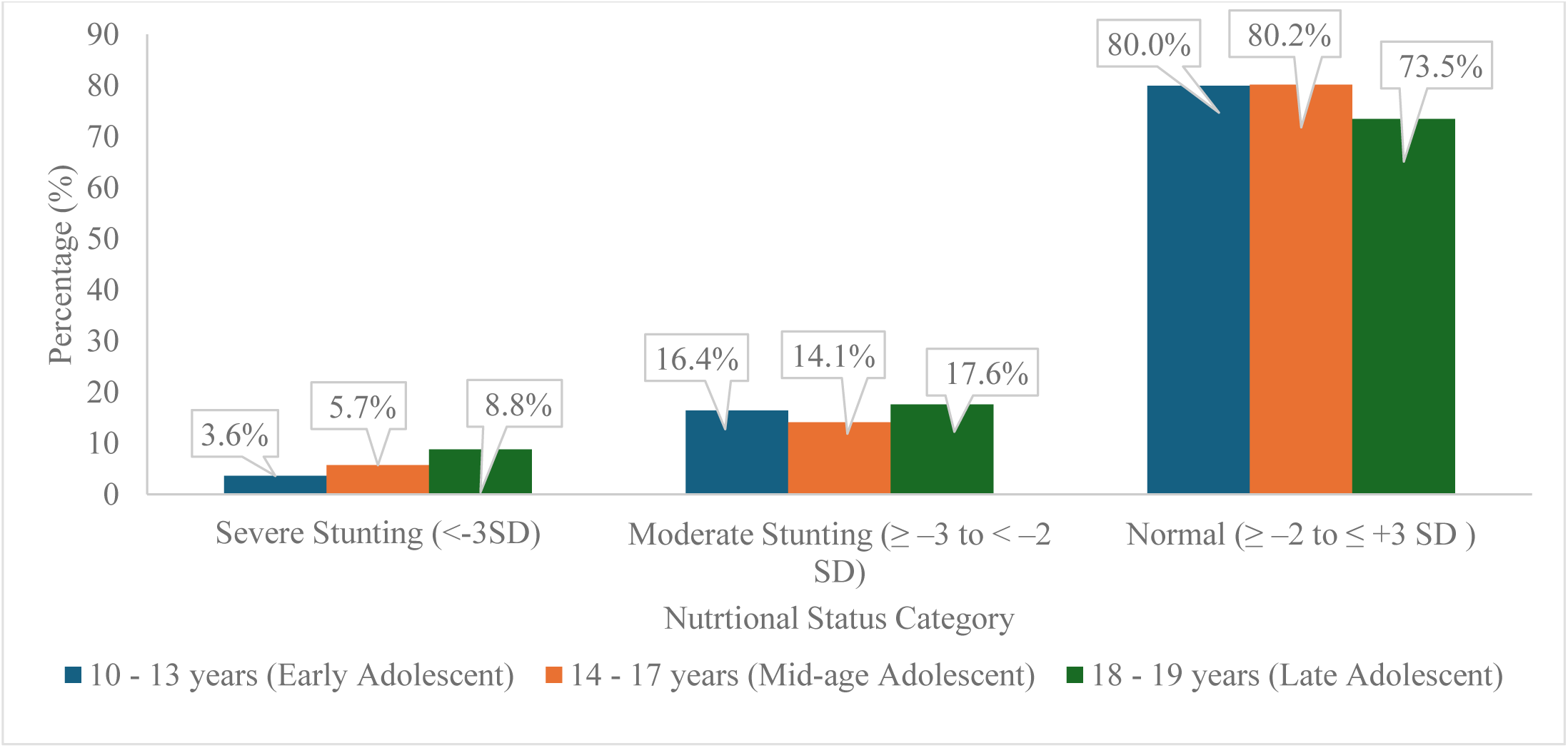
Distribution of stunting status related to age category of ALHIV on ART follow-up.

### **c)** Acute Malnutrition Based on MUAC-for-Age

Based on MUAC-for-age criteria, 34.9% (n = 134) of participants were malnourished ^[28]^, including 80.6% (n = 108) with moderate acute malnutrition and 19.4% (n = 26) with severe acute malnutrition.

Mid-adolescents (ages 14–17) had the highest prevalence of acute malnutrition (37.4%, n = 85), including 8.4% (n = 19) with severe acute malnutrition. In comparison, the early adolescent group had a lower prevalence of severe acute malnutrition (3.6%, n = 2), as did late adolescents (4.9%, n = 5). Moderate acute malnutrition was most prevalent among early adolescents (34.5%) (Figure 3a).

**Figure 3a.**
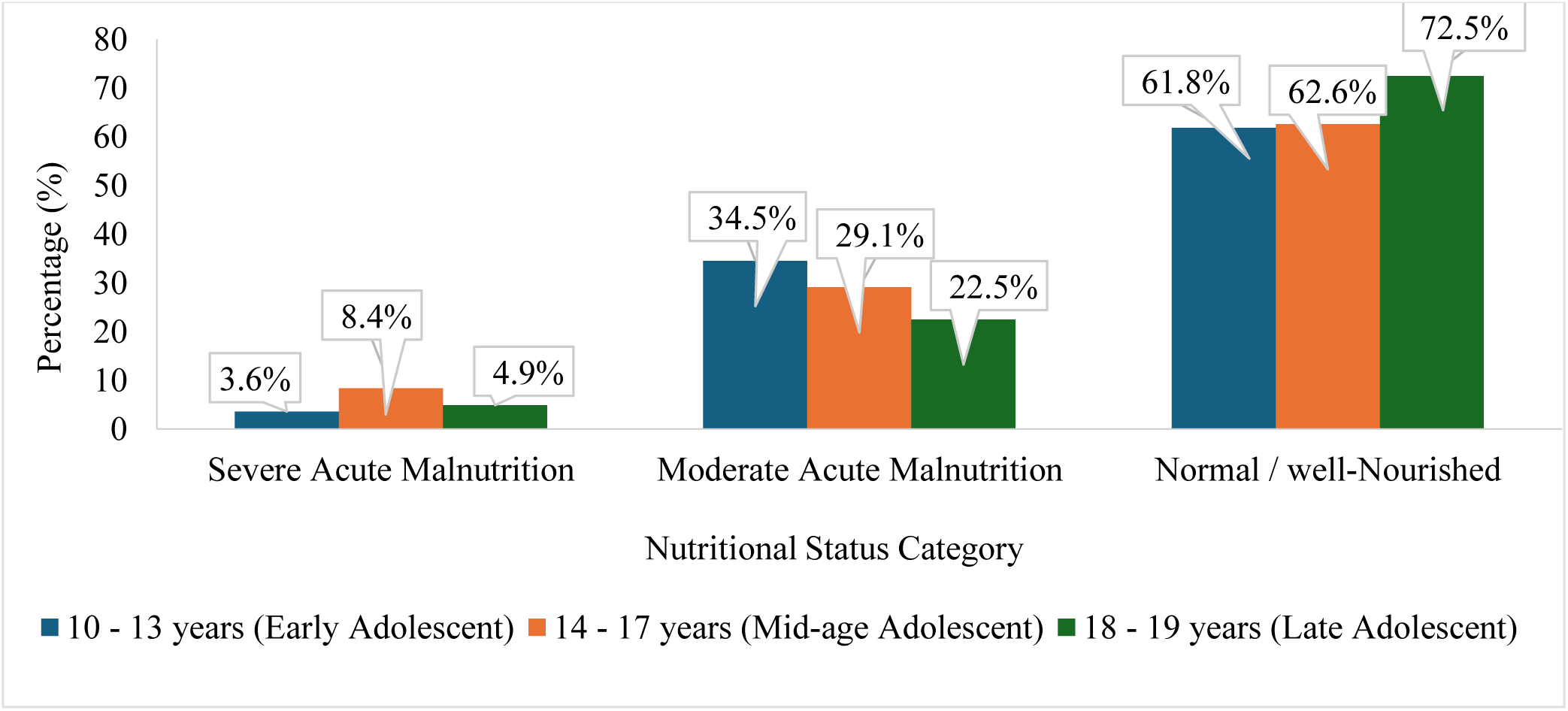
Distribution of malnutrition status according to MUAC measurement by age category for ALHIV on ART follow-up

Male participants were more likely to be acutely malnourished (41.2%, n = 73) compared to females (29.4%, n = 61). Severe acute malnutrition was recorded in 9.6% of males (n = 17) versus 4.3% of females (n = 9) (Figure 3b).

**Figure 3b.**
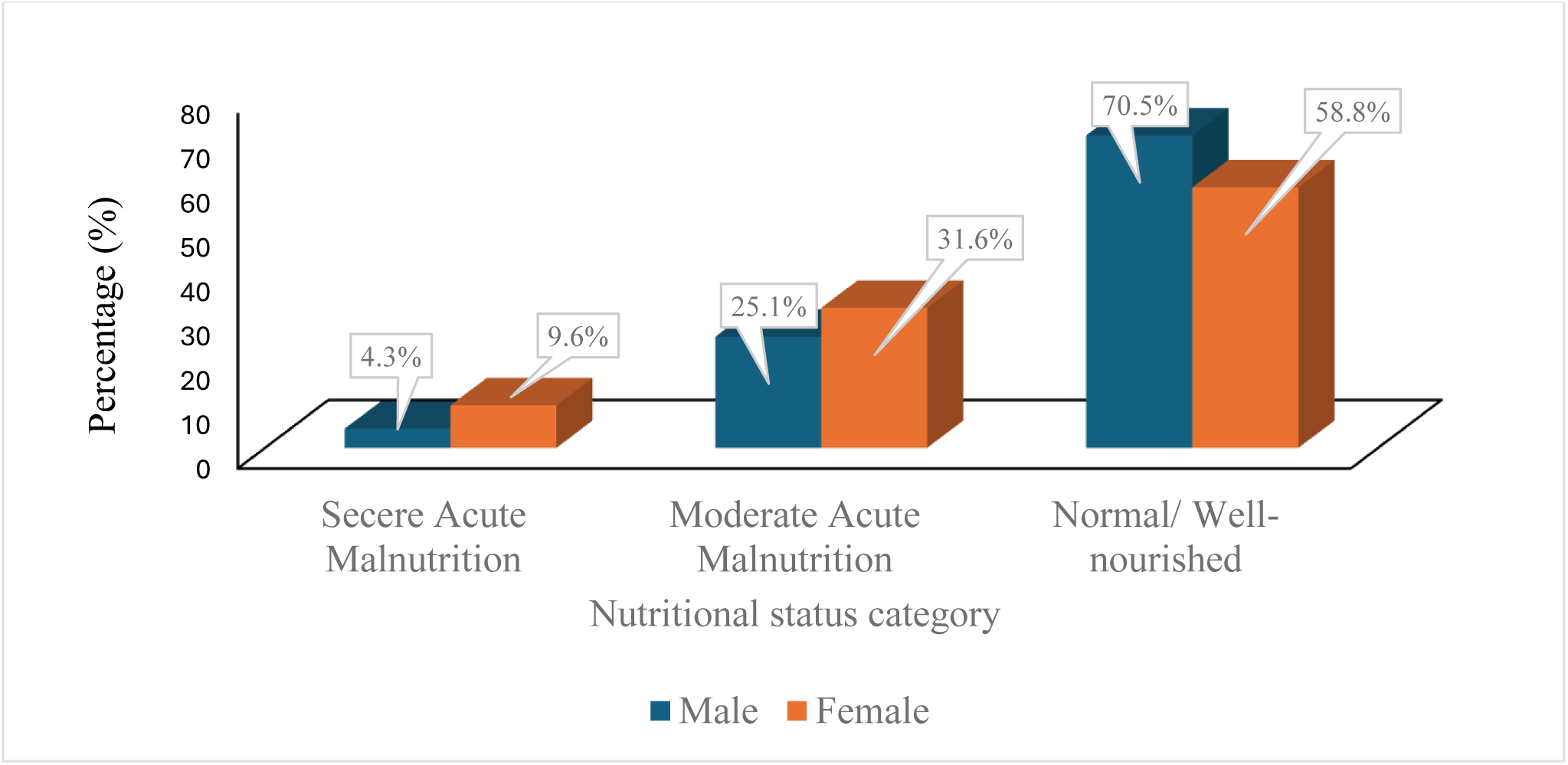
Distribution of malnutrition status according to MUAC measurement by gender for ALHIV on ART follow-up

### 3.3. Anthropometric parameters associated with malnutrition

#### 3.3.1. Correlation analysis

Moderate positive correlations were found between the sum of four skinfold thickness measurements and both body fat percentage (r = 0.436, p < 0.001) and BMI-for-age (r = 0.335, p < 0.001), suggesting that increases in skinfold thickness are associated with higher adiposity and BMI. The correlation between skinfold thickness and MUAC-for-age was weaker (r = 0.296, p < 0.001), indicating that MUAC may reflect muscle mass and other components not directly captured by skinfold measures (Table 5).

**Table 5a.** Nutritional assessment practices reported by clinic healthcare workers for attendees at the ART clinics of selected hospitals in two regions of Ethiopia, 2024.

| Variables | Description | Frequency<br>N (%) |
| --- | --- | --- |
| Anthropometric measurements used in ART clinics | Height | 44 (100) |
|  | Weight | 44 (100) |
|  | Body Mass Index | 42 (95.5) |
|  | Mid-upper Arm Circumference | 25 (56.8) |
|  | Skinfold Thickness | 0 (0.0) |
|  | Waist Circumference | 3 (6.8) |
|  | Waist-to-hip ratio | 2 (4.5) |
|  | Lean body mass | 0 (0.0) |
|  | Body Fat | 0 (0.0) |
|  | Grip strength | 0 (0.0) |
| Biochemical Assessments undertaken for ALHIV in ART clinics | Glucose Test | 32 (72.7) |
|  | Blood Urea and Nitrogen test | 33 (75.0) |
|  | Creatinine Test | 32 (72.7) |
|  | BUN to Creatinine Ratio | 16 (36.4) |
|  | Calcium | 4 (9.1) |
|  | Total Protein | 3 (6.8) |
|  | Albumin | 20 (45.5) |
|  | Alkaline Phosphatase | 21(47.7) |
|  | Alanine Aminotransferase | 22 (50.0) |
|  | White Blood Cell count | 35 (79.5) |
|  | Red Blood Cell count | 39 (88.6) |
|  | Hemoglobin | 39 (88.6) |
|  | Hematocrit | 39 (88.6) |
|  | Mean corpuscular Volume | 38 (86.4) |
|  | Mean Corpuscular Hemoglobin | 38 (86.4) |
|  | Platelet count | 25 (56.8) |
|  | Helminth Infection (hookworm and ascaris) | 17 (38.6) |
| Clinical Assessment conducted at the ART clinic | Bilateral pitting oedema | 40 (90.9) |
|  | Visible wasting | 41 (93.2) |
|  | Recent weight loss | 41 (93.2) |
|  | Dermatosis | 18 (40.9) |
|  | Eye signs (Bitot spot, corneal cloudiness, conjunctivitis, corneal ulceration) | 17 (38.6) |
|  | Palms, mucus membranes, and nail bed loss | 35 (79.5) |

**Table 5b.** Nutritional counselling and education practices reported by clinic healthcare workers for attendees at the ART clinics of selected hospitals in two regions of Ethiopia, 2024.

| **Variables** | **Frequency**<br>**N (%)** |
| --- | --- |
| Availability of nutrition counselling and education for ALHIV at ART clinic (n=44) | 17 (38.6) |
| Regular practice of nutritional counselling and education to ALHIV during ART follow-up (n=17) | 10 (58.8) |
| Use GALIDRAA approaches nutrition counselling and education for ALHIV (n=17) | 4 (23.5) |
| Use ORPA approaches during discussion time for nutrition counselling and education for ALHIV (n=17) | 2 (11.8) |

ALHIV – Adolescent Living with HIV; GALIDRAA –Greet, Ask, Listen, Identify, Discuss, Recommend, Agree, and Appoint; ORPA –Observe, Reflect, Personalize, and Act approaches during discussion time for nutrition counselling for ALHIV

**Table 5c.** Nutritional management and care/support practices of the study participants in selected hospitals of two regions, Ethiopia, 2024 (n=44)

| Variables | Description | Frequency<br>N (%) |
| --- | --- | --- |
| Has standard admission criteria for nutrition care and support | Yes | 21 (47.7) |
| Types of standard admission criteria used (n=21) | Anthropometric Measurement (i.e., only BMI < 18.5kg/m<sup>2</sup>) | 17 (80.9) |
|  | Combined Method (i.e., Anthropometric* and Clinical findings**) | 4 (19.1) |
| Nutrition screening | Yes | 36 (81.8) |
| Nutrition Case Management | Yes | 36 (81.8) |
| Nutrition Supplementation | Yes | 32 (72.7) |
| Type of supplementation provided [multiple responses] (n=32) | Ready to use Therapeutic Food (RUTF) | 32 (100) |
|  | Ready to use Supplementary Food (RUSF) | 5 (15.6) |
|  | Others*** | 2 (6.25) |
| Supplementation delivery period (n=32) | Every 2 weeks | 5 (15.6) |
|  | Every month | 27 (84.4) |
| Availability of a strategy to detect sharing supplementation e.g. with a sibling (n=32) | Yes | 13(40.6) |
| Strategies used to detect sharing (n=13) | Progress Assessment | 3 (23.1) |
|  | Weight change/gain assessment | 10 (76.9) |
| Has standard discharge criteria for nutrition care and support program | Yes | 16 (36.4) |

N.B: \*Anthropometric measurements like MUAC<16cm, BMI<18.5kg/m<sup>2</sup>, Weight-for-height Z-
Score < -2 or -3
\*\*Clinical signs like Visible severe wasting, recent weight loss, Bilateral pitting Oedema
\*\*\*Others – Often supplementation of Oil, Teff flour, and Wheat flour

#### 3.2.2. Regression Analysis

Simple linear regression identified significant associations between several anthropometric measures and thinness, as defined by BMI-for-age indices. These included:

- MUAC < 21 cm (β =-0.38)
- Skinfold thickness (S4SKT) < 25 cm (β =-0.34)
- Hip circumference < 82 cm (β =-0.33)
- Waist circumference < 64 cm (β =-0.21) (Table 6)

**Table 6.**
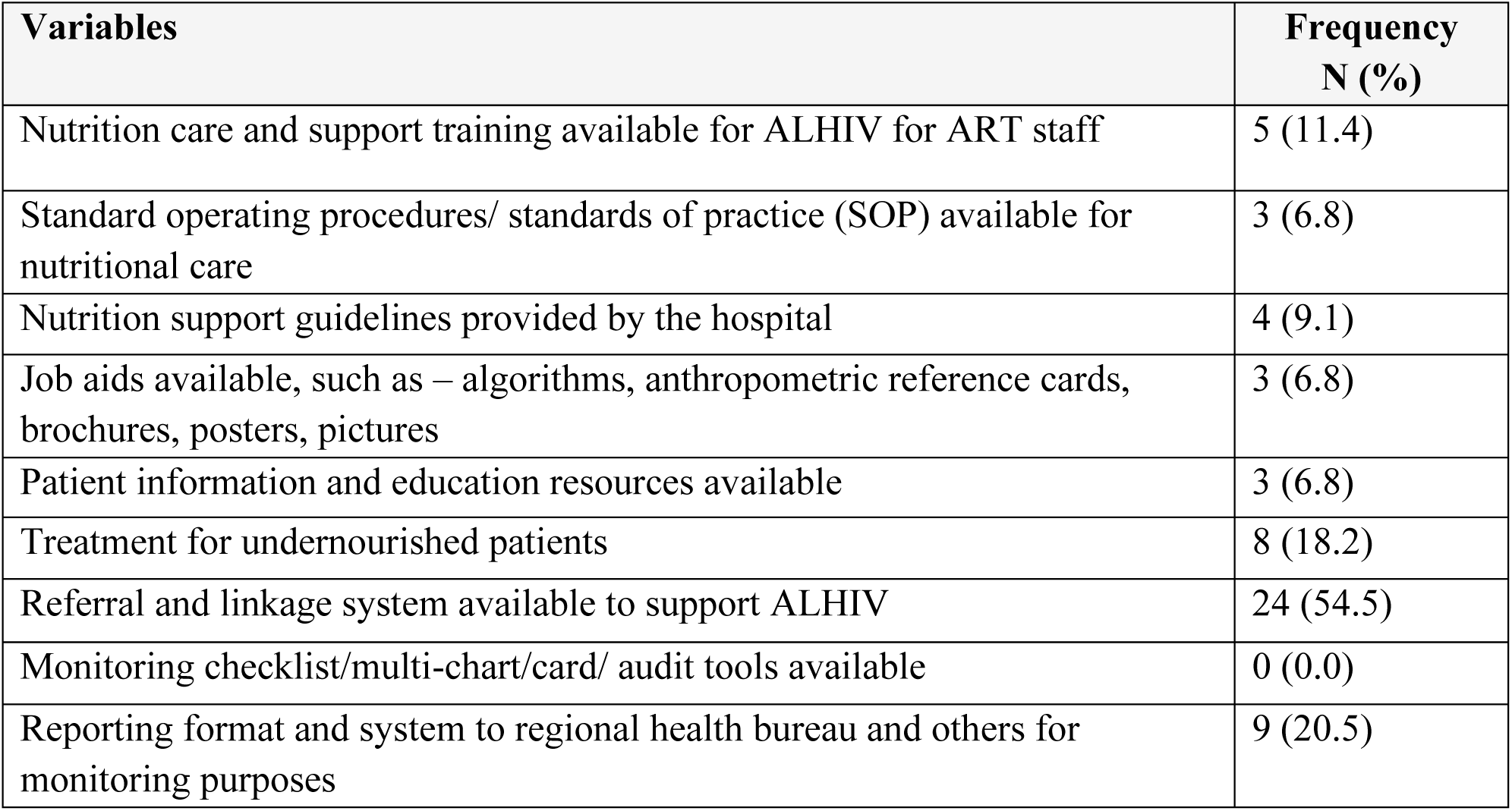
Resources and activities to support staff in providing nutritional care and support for ALHIV on ART follow-up (n=44)

| **Variables** | **Frequency**<br>**N (%)** |
| --- | --- |
| Nutrition care and support training available for ALHIV for ART staff | 5 (11.4) |
| Standard operating procedures/ standards of practice (SOP) available for nutritional care | 3 (6.8) |
| Nutrition support guidelines provided by the hospital | 4 (9.1) |
| Job aids available, such as – algorithms, anthropometric reference cards, brochures, posters, pictures | 3 (6.8) |
| Patient information and education resources available | 3 (6.8) |
| Treatment for undernourished patients | 8 (18.2) |
| Referral and linkage system available to support ALHIV | 24 (54.5) |
| Monitoring checklist/multi-chart/card/ audit tools available | 0 (0.0) |
| Reporting format and system to regional health bureau and others for monitoring purposes | 9 (20.5) |

Multivariable stepwise backward regression confirmed these associations. A MUAC < 21 cm was independently associated with lower BMI-for-age Z-scores (β =-0.37, 95% CI:-0.42 to-0.26). A S4SKT < 25 cm was also significantly associated with thinness (β =-0.25, 95% CI:-0.29 to-0.13). Additionally, hip circumference < 82 cm was linked to a β of-0.16 (95% CI:-0.23 to-0.06).

These findings highlight the value of MUAC, skinfold thickness, and hip circumference as practical anthropometric indicators for identifying ALHIV at risk of malnutrition, reinforcing the need for routine monitoring and targeted nutritional interventions.

### 3.4. Healthcare Professionals’ Nutritional Assessment, Counselling, and Support Practices

#### 3.4.1. Nutritional Assessment Practices

Most healthcare professionals (HCWs) reported using multiple methods to assess the nutritional status of ALHIV. Commonly utilized methods included anthropometric measurements (100%, n = 44), clinical assessments (93.3%, n = 41), and biochemical assessments (77.3%, n = 34). However, dietary intake assessments and food security evaluations were less commonly conducted, reported by only 36.4% (n = 16) and 27.3% (n = 12) of participants, respectively. Only one-quarter (25%, n = 11) reported using all four assessment methods (Table 7).

All HCWs reported access to standardized weight and height measurement tools. However, only 63.6% (n = 28) used MUAC tapes, 9.1% (n = 4) had access to waist circumference tapes, and just 4.5% (n = 2) reported using skinfold calipers. No participants had access to more advanced tools such as dual-energy X-ray absorptiometry (DXA) or grip strength dynamometers.

Weight-for-age, height-for-age, and weight-for-height indices were routinely assessed by all HCWs. Most also reported regular evaluation of BMI (95.5%, n = 42), and more than half assessed MUAC (56.8%, n = 25). However, few assessed waist circumference or waist-to-hip ratio. None conducted comprehensive body composition assessments (e.g., skinfold thickness, lean body mass, or grip strength).

While biochemical testing was reported as routine practice, specific markers such as total protein and calcium were not frequently assessed. Although visual inspection for signs of malnutrition was commonly performed, less than half of HCWs routinely assessed for dermatosis or ocular signs of micronutrient deficiency (Table 7).

#### 3.4.2. Counselling and Nutrition Education

Only 38.6% (n = 17) of HCWs reported providing nutrition counselling and education during routine clinic visits, and among these, just over half (58.8%, n = 10) offered these services regularly. The WHO-endorsed GALIDRAA counselling framework was used by 23.5% (n = 4), while only 11.8% (n = 2) implemented the ORPA strategy during the discussion phase of counselling (Table 8).

#### 3.4.3. Nutrition Care and Support Services

Approximately half of the participants (47.7%, n = 21) reported the presence of admission criteria for nutrition care services within their clinics, while only 36.4% (n = 16) indicated the presence of discharge criteria. Most clinics used BMI < 18.5 kg/m² as the sole admission threshold for nutritional care (81%, n = 17).

Nutrition screening (81.8%, n = 36), case management (81.8%, n = 36), and supplementation (72.7%, n = 32) were reported by the majority of respondents. All undernourished ALHIV were provided with ready-to-use therapeutic food (RUTF), though only 15.6% (n = 5) received ready-to-use supplementary food (RUSF). Most healthcare professionals (84%, n = 27) provided monthly supplementation; however, only 40.6% had strategies in place to monitor or prevent the sharing of supplements among siblings (Table 9).

#### 3.4.4. Implementation Support

Most participants reported limited institutional support for nutritional care in ART clinics. Key gaps included the absence of standard operating procedures (SOPs), national nutrition guidelines, educational materials, and monitoring or reporting tools. Few participants had access to staff training, or on-site treatment services for undernourished ALHIV. Nonetheless, more than half (54.5%, n = 24) reported access to external referral systems for additional support (Table 10).

#### 3.4.5. Healthcare Professionals’ Perspectives

Over half of the respondents (54.5%, n = 24) expressed dissatisfaction with current nutrition assessment, counselling, and care practices for ALHIV. One participant noted:

*“Nutrition assessment, counselling, care, and support services for adolescents living with HIV are inadequate. This perception stems from a lack of integration of nutrition services and insufficient training for ART clinic staff on HIV and nutrition.”*

Despite routine reporting of anthropometric assessments, only 36.4% (n = 16) consistently conducted all three key measurements: weight, height, and BMI. Moreover, just 13.6% (n = 6) reported conducting effective clinical assessments, such as dietary intake evaluation or identifying clinical signs of malnutrition.

A respondent highlighted the complexity of nutritional assessment in practice:

*“Nutritional assessments are crucial, but limited methods are relied upon in practice. Given the complexity of malnutrition, accurately determining nutritional status is challenging.”*

Only 11.4% (n = 5) reported that nutrition supplementation and monitoring services were effectively delivered, and even fewer (4.5%, n = 2) noted effective delivery of psychosocial support.

Overall, 34% (n = 15) of healthcare professionals were dissatisfied with the quality of nutrition services at their facilities, citing the lack of integration with HIV care, incomplete screening protocols, insufficient training, and the absence of consistent nutritional support programs.

## 4. Discussion

This study reveals a critical public health concern: high levels of malnutrition among adolescents living with HIV (ALHIV) in Ethiopia. Utilizing multiple anthropometric indicators—BMI-for-age (BAZ), height-for-age (HAZ), and mid-upper arm circumference (MUAC)—we found alarmingly high rates of thinness, stunting, and acute malnutrition, particularly among mid-adolescents and male participants. These findings underscore the urgent need for targeted, age-and gender-specific nutritional interventions.

The observed prevalence of malnutrition aligns with studies from Uganda and southern Ethiopia ^[14, 29]^, yet exceeds rates reported in southwest Nigeria ^[30]^ and other regions of Uganda ^[31]^. Such variation may reflect differences in socioeconomic status, dietary practices, healthcare access, and the availability of nutrition-focused services. The underutilization of structured and evidence-based nutritional interventions further compounds these disparities.

Despite regular basic anthropometric measurements in ART clinics, the absence of more sensitive assessments—such as skinfold thickness, waist circumference, and DXA—limits healthcare providers’ ability to detect early or subclinical malnutrition. This shortfall likely contributes to the high burden of malnutrition observed in this cohort.

Furthermore, our findings reveal a substantial gap in nutrition counselling and education. While a minority of healthcare workers provide nutrition education, the limited use of structured tools such as GALIDRAA and ORPA indicates a lack of training and institutional support. These frameworks are crucial for delivering personalized, high-quality care, and their absence is likely to reduce the effectiveness of counselling practices.

Although most healthcare providers conduct nutrition screening and provide RUTF to severely malnourished ALHIV, many report inadequate access to assessment tools, treatment guidelines, and support services. Consistent with previous research ^[32, 33]^, this lack of resources and training undermines the ability of clinics to deliver integrated, sustained nutritional care. The infrequent use of ready-to-use supplementary foods (RUSF) for moderate malnutrition and the lack of psychosocial support services further highlight systemic gaps in care delivery.

Healthcare workers’ perspectives reinforce these findings. Over half expressed dissatisfaction with existing nutrition services, citing poor integration with HIV care, absence of comprehensive screening, and limited access to tools and training. These concerns reflect an urgent need for investment in standardized protocols, staff development, and sustainable service delivery.

The heightened vulnerability of mid-adolescent males to malnutrition highlights the importance of age-and sex-specific nutritional interventions. While more than half of healthcare workers had access to referral and linkage systems, better integration of nutrition into ART care, along with improved capacity building, is essential to improving outcomes. Addressing these issues requires a multi-faceted response including training, guideline development, structured assessment protocols, and consistent supplementation strategies.

### 4.1. Implications of the Study

This study offers several important implications for strengthening nutritional care for ALHIV in Ethiopia:

- **Clinical Practice**: Healthcare providers should incorporate more sensitive and diverse nutritional assessment techniques. This requires targeted training and improved access to equipment.
- **Counselling and Education**: Evidence-based counselling approaches (e.g., GALIDRAA, ORPA) should be embedded into routine care, supported by ongoing education and mentorship for healthcare workers.
- **Public Health and Policy**: Socioeconomic barriers to nutrition should be addressed through community-based food security initiatives. National HIV policies should include standardized nutritional protocols, backed by adequate funding and logistical support.
- **Research**: Future research should explore effective, context-specific nutritional interventions for ALHIV, integrating mixed-methods and longitudinal designs to assess impact over time.

### 4.2. Limitations

This study has several limitations:

- **Self-reported data** from ALHIV and healthcare providers may be subject to recall and social desirability bias, potentially affecting the accuracy of findings. Although steps were taken to minimize bias through tool validation and triangulation, residual inaccuracies may remain.
- **Sample size** for the healthcare worker survey was modest, limiting generalizability. The small number of qualitative responses may not fully capture the range of provider perspectives.
- **Variability in training** among data collectors may have affected the consistency of measurements, and some dietary recall data may have been incomplete or imprecise.
- **Cross-sectional design** restricts inference of causal relationships. Longitudinal or interventional studies are needed to better assess the effectiveness of nutrition interventions over time.
- **External factors** such as socio-political instability, regional resource allocation, and unmeasured institutional differences may have influenced findings and were not controlled for in this study.

Future research should incorporate larger and more diverse samples, objective nutrition indicators, and longitudinal approaches to strengthen evidence on effective nutritional support for ALHIV.

## 5. Conclusion and recommendations

This study highlights the urgent need for comprehensive, integrated nutritional care for adolescents living with HIV in Ethiopia. Although basic assessment and support practices are present in many ART clinics, the underuse of sensitive assessment tools and structured counselling frameworks limits their effectiveness. Targeted training, standardized national guidelines, and improved resource allocation are essential to strengthening nutritional services.

Implementing age-and gender-specific interventions, improving assessment protocols, and integrating nutrition counselling into HIV care can help reduce the burden of malnutrition among ALHIV. Future studies should include larger, multi-site samples and longitudinal methodologies to assess the long-term impact of nutritional interventions. Scaling up support for healthcare workers and ensuring consistent access to assessment tools and therapeutic foods are crucial next steps to ensure that ALHIV receive the comprehensive care they need to thrive.

## Declaration

### Authors contribution

MGB performed data collection, analysis and drafted the initial manuscript. All authors contributed to the conceptualization, methodology development, data analysis, interpretation, revision and approval of the final manuscript.

## Data Availability

The data supporting the findings of this study are available upon reasonable request. Access to the data may be restricted to ensure participant confidentiality and to comply with ethical guidelines.

## Acknowledgments

The authors wish to acknowledge the Local Government Areas of Ethiopia, the Health Bureau, and all hospital administrators for granting permission to collect data. We also extend our gratitude to the ART unit staff members and patients for their participation in the study.

## Supplementary Files

### Supplementary File Box 1: Clinical Measurements Protocol

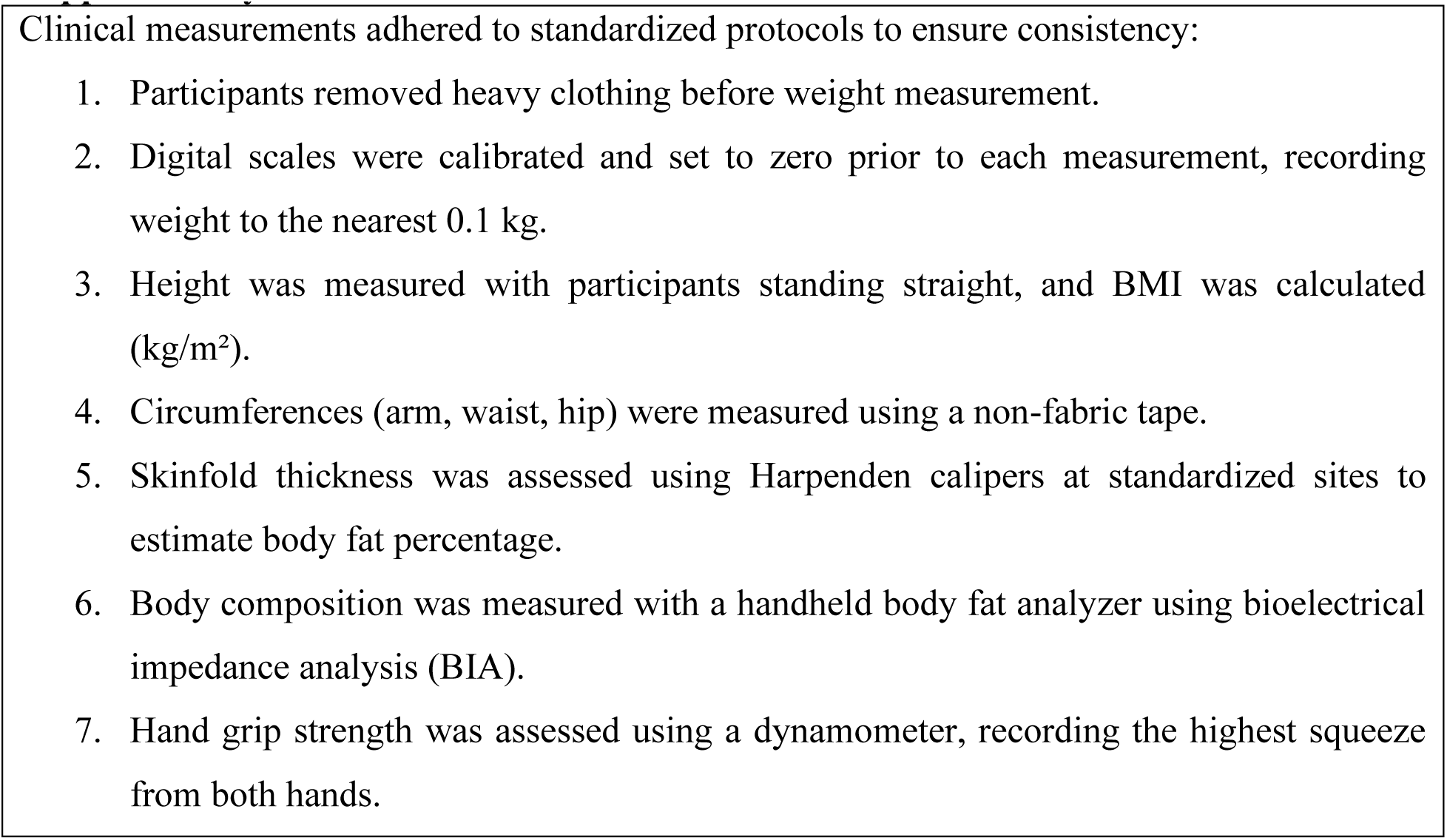

### Supplementary File Box 2: Operational definition

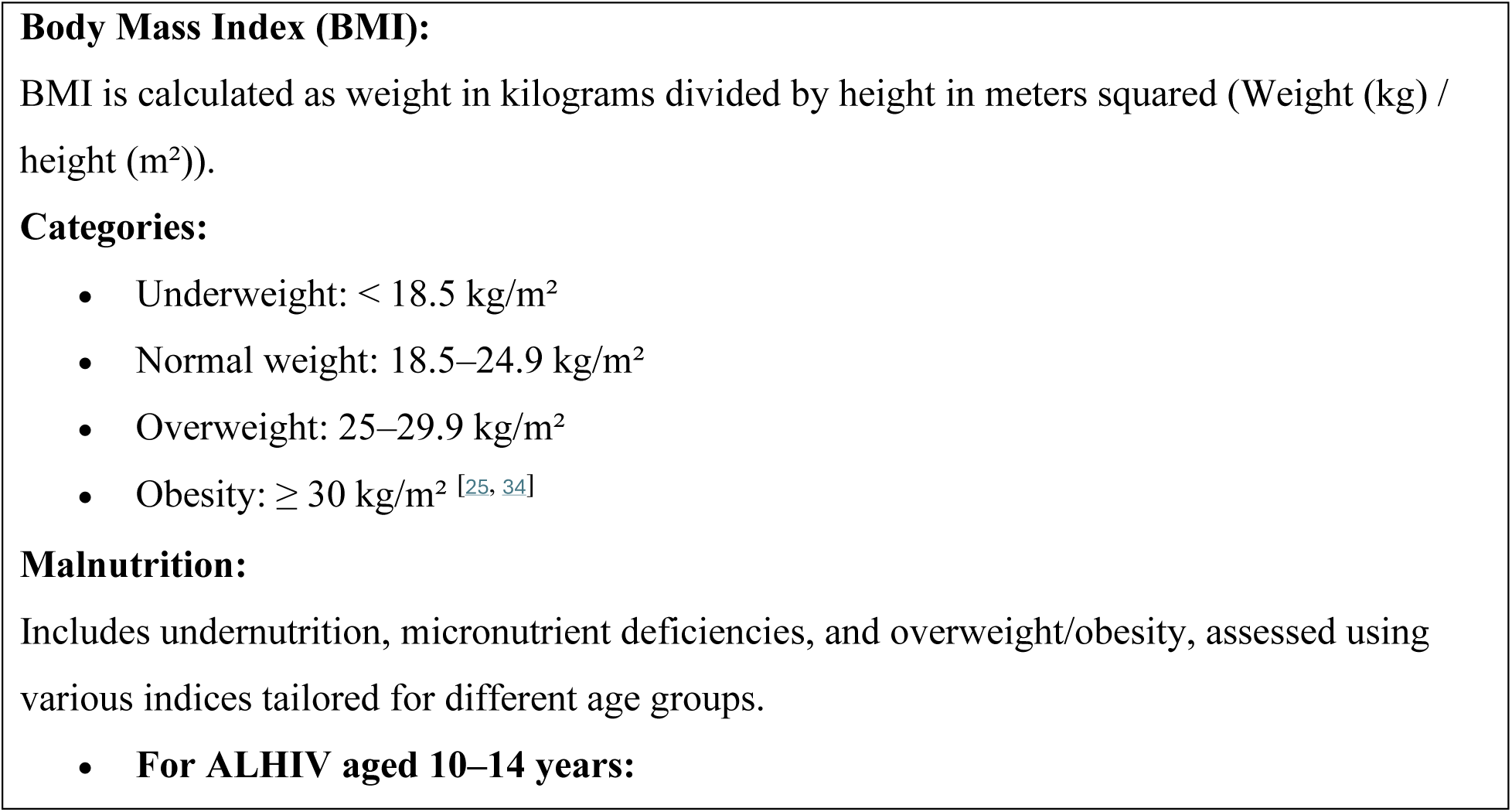
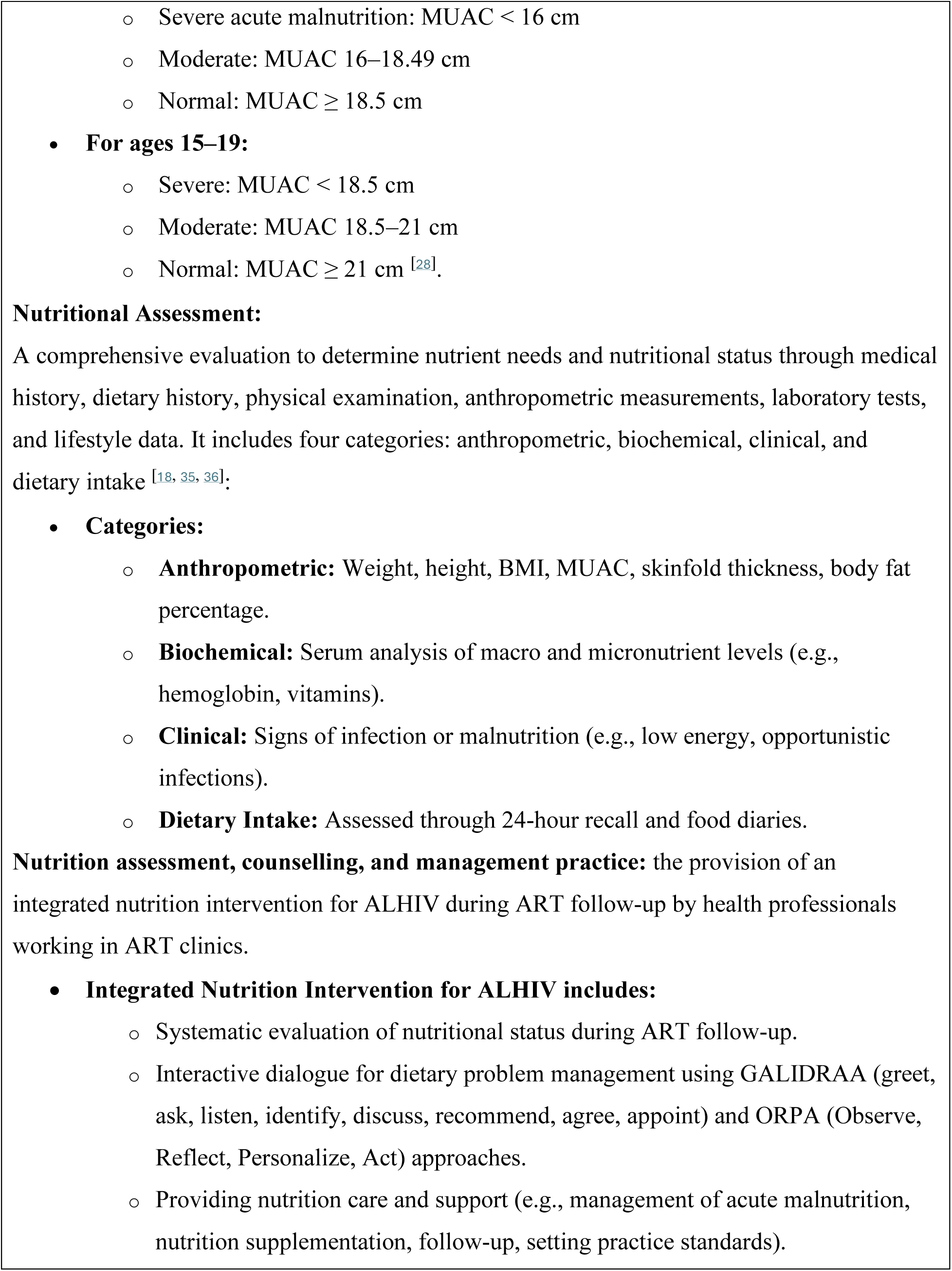
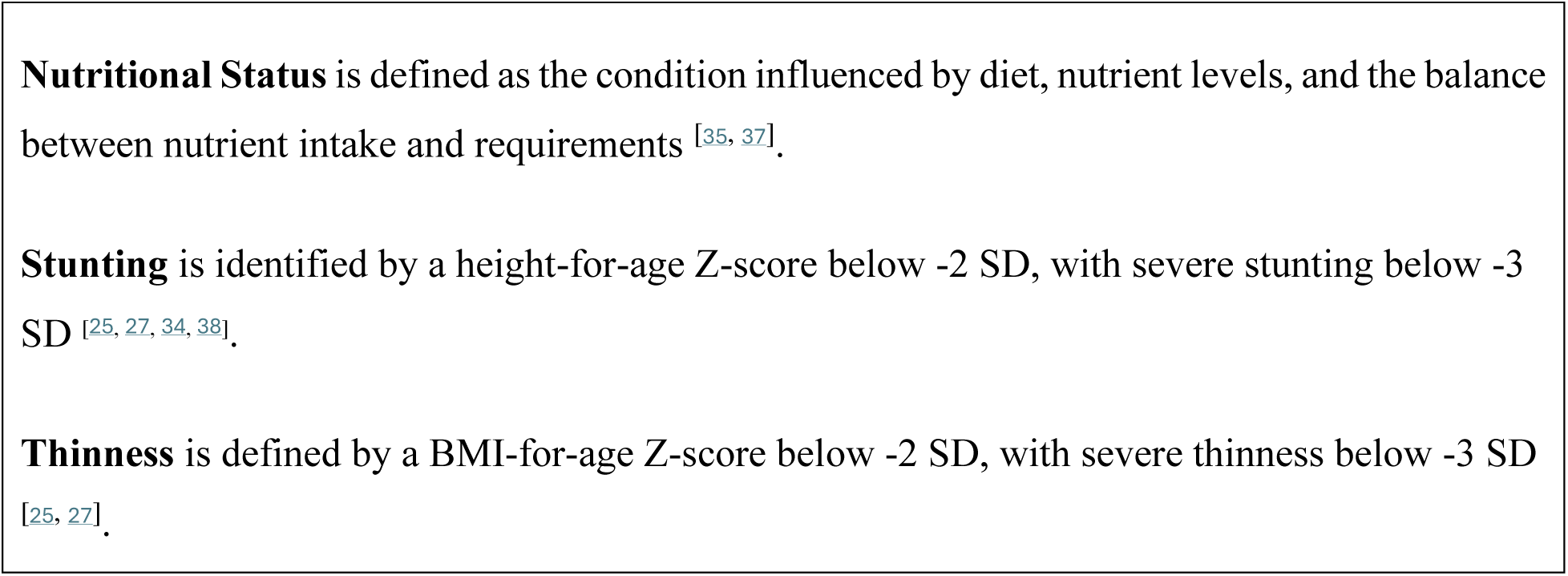

